# At home and online during the early months of the COVID-19 pandemic and the relationship to alcohol consumption in a national sample of U.S. adults

**DOI:** 10.1101/2020.09.20.20197608

**Authors:** Karen G. Chartier, Jeanine P.D. Guidry, Catherine A. Lee

## Abstract

**Objective:** The current study seeks to understand the links between social media use and alcohol consumption during the early months of the COVID-19 pandemic.

**Method:** Data were from the national Understanding American Study, a probability-based Internet panel weighted to represent the U.S. population. Subjects (N=5874; 51% female) were adults, 18 years and older, who completed a March survey (wave 1) and a follow-up survey one month later (wave 3). Analyses assessed the relationship of social media use at wave 1 with wave 3 alcohol use, accounting for wave 1 alcohol use and the sociodemographic characteristics of the sample. We examined the effect of working or studying from home as a moderator.

**Results:** Twitter and Instagram users, but not Facebook users, drank more frequently at wave 3 than non-users. For Instagram users, more frequent alcohol use at wave 3 was at least partially attributed to the frequency of drinking at wave 1. The interaction between Twitter use and working or studying from home was statistically significant. The combination of being on Twitter and working or studying from home was associated with drinking more days a week.

**Conclusions:** Exposure to content about COVID-19 and increased alcohol consumption during the pandemic may contribute to more frequent alcohol use for some social media users, especially those sheltering at home. The study of public health messaging via social media to change alcohol use behaviors during traumatic events is warranted.

## Introduction

As the effects of COVID-19, beyond the virus itself, begin to emerge, the U.S. is seeing the economic, social, and psychological tolls, some of which became apparent immediately but many of which may emerge over time. Months into the response, we are observing a convergence of behaviors that present some worrisome trends. In Australia, individuals who lost their jobs during the early months of the pandemic and who reported higher levels of stress, depression, sleep changes, and overeating had a greater chance of increased alcohol consumption (1). A survey in Europe showed that more than 30% of participants changed their drinking habits during the early months of the COVID-19 pandemic, with some reporting lower and others higher alcohol consumption (2). While there was an initial suggestion that challenged supply chains and store closures would result in a shortage of alcohol, many alcohol distributors and delivery services have become creative to ensure that scarcity is not an issue (3). In fact, there was a spike in wine and spirit sales as fears of COVID-19 spread (4). There was even misinformation being spread that alcohol ingestion could help prevent or cure the COVID-19, which was quickly discounted by the National Institutes of Health (NIH) and the World Health Organization (WHO) (5,6)

With the emergence of working from home for many populations, and government and medical mandates on social distancing, home electronic usage, and more specifically social media usage, has increased (7–9). Some of the increase in social media use has been a method for people to stay up to date on COVID-19 and other news events (9). Social media use has also been described as a form of “escapism,” to alleviate stress and avoid difficult or problematic thoughts (10). Alternatively, studies have also shown social media to have positive healthcare implications by providing access to public health information and encouraging peer and emotional support (11).

However, the WHO (12) has warned of an “infodemic” in the age of COVID-19, described as “an over-abundance of information–some accurate and some not–that makes it hard for people to find trustworthy sources and reliable guidance when they need it”. A recent study among a Chinese population showed that this increase in social media exposure during the COVID-19 outbreak correlated with an increase in anxiety and depression (13). In addition, alcohol-related content has long been prevalent on social media platforms (14), as evident by the trending hashtag #quarantini during the early days of the pandemic (15). Prior studies suggest that both posting of and exposure to alcohol-focused content on social media is associated with higher rates of consumption and cravings, as well as clinical disorders (14,16,17).

Prior studies further show that traumatic events, stress, and alcohol consumption are correlated. In a review article, Keyes et al. (18) reported that exposure to disastrous or traumatic events, whether manmade or natural, leads to an increase in overall alcohol consumption for those experiencing the stress, as well as serving as a coping mechanism for individuals with a history of alcohol use disorder. Multiple factors are related to the level of increased consumption, including the severity and expectedness of the trauma, whether it is physical or emotional trauma, and the origin of the stressor (18). Wiederhold (9) further suggested that in scenarios involving trauma or disaster, the number of fatalities will often be significantly outnumbered by the survivors who experience negative mental health outcomes as a result of the stress and trauma related to the disaster. Because of COVID-19’s recentness, determining the interplay of these factors related to COVID-19 as a traumatic event and the longitudinal effects, to include alcohol consumption, should be studied at a later date.

For the current study, we examine the relationship between alcohol consumption and social media use during the early months of the COVID-19 pandemic. As this review of relevant studies shows, there has been plentiful research on social media exposure and alcohol use and on traumatic events and alcohol use. What we know far less about is the relationship between social media use and alcohol consumption during a traumatic event. We further examine the effect of working or studying from home as a moderator of this relationship, given its association with increased social media exposure. Concern was raised early on that the sudden isolation of millions would be associated with increased alcohol use (19). Drawing from the reviewed areas of research and the recent study of social media exposure and anxiety and depression (13), we expect that social media users will consume more alcohol during the COVID-19 pandemic, and that the positive relationship between social media use and alcohol consumption will be exacerbated for individuals who work or study from home. While most studies examining social media and alcohol use have been conducted in adolescents and college students or young adults (14), we extend this work by testing these relationships in an adult general population sample in the U.S.

## Methods

### Sample

The current sample (*N*=5874) from the national Understanding American Study (UAS) included adults, 18 years and older. Begun in 2014, UAS is an ongoing, probability-based internet panel, with members recruited via address-based sample waves of all U.S. households, with internet-connected tablets provided to households not previously online (20). A total of 6,933 panel members from the U.S.-weighted sample participated in a March 2020 survey, and then on April 1, 2020, were invited to participate in an ongoing coronavirus tracking survey. For the current study, we included those weighted sample members who completed both the wave 1 survey between March 10 and 31 and the wave 3 survey between April 15 and 30. We did not include responses from the wave 2 survey, collected between April 1 and 14, as we sought to examine alcohol use across differentiated time frames. Surveys were completed online using a computer, mobile phone, or tablet and in English or Spanish according to the respondent’s preference. The participation rate was 82% in March for the weighted sample, and approximately 96% of respondents completed their wave 3 survey in April (20,21). Data are weighted to account for the sampling procedure and probabilities of selection and to be representative of the U.S. population, as described elsewhere in more detail (20,21).

Table 1 presents the sociodemographic characteristics of the sample. About half of respondents were female, and the majority were non-Hispanic White, married, had at least some college education, were employed, and 40 years of age or older. The average number of household members was slightly under two. These sample characteristics are comparable with census estimates in regards to sex, but include a somewhat larger percentage than in the U.S. population of non-Hispanic Whites, persons with a bachelor’s degree, and those 65 years and older, respectively, 60.1%, 31.5%, and 16.5% (22). The number of individuals per household is somewhat smaller in the current sample than the U.S. population estimate, which was 2.63 (22).

**Table 1.** Sample socio-demographics and social media use (*N*=5874)

|  | **% or *M* (SE)** |
| --- | --- |
| **Sex (% Female)** | 50.91 |
| **Age (%)** |  |
| 18-29 years | 12.20 |
| 30-44 years | 32.11 |
| 45-64 years | 34.86 |
| 65 years or older | 20.82 |
| **Race/ethnicity (%)** |  |
| Asian* | 5.25 |
| Black* | 11.53 |
| Hispanic/ Latino | 15.35 |
| White* | 64.44 |
| Other groups* | 3.43 |
| **Education (%)** |  |
| Less than high school | 8.45 |
| High school/ GED | 28.93 |
| Some college | 27.96 |
| Bachelor's degree + | 34.66 |
| **Currently employed (% Yes)** | 60.77 |
| **Marital status (%)** |  |
| Married | 56.72 |
| Previously married** | 19.26 |
| Never married | 24.01 |
| **Household members, *M* (*SE*)** | 1.78 (.028) |

*Notes:* *M* = mean; *SE* = standard error; GED = general educational diploma; \*non-Hispanic; \*\*separated/divorced/widowed

## Measures

### Social media use

was collected at wave 1. Respondents indicated whether they used Twitter, Instagram, and/or Facebook, with each item coded 0 no and 1 yes. Affirmative responses indicated that respondents had an account and used it. Those indicating that they had an account, but never used it, and those who did not have an account were coded no. Active users reported their average minutes on social media in a day across platforms. Responses over 1440 minutes were re-coded to 1440 to indicate near constant social media use, and respondents reporting no social media use had zero minutes. Additionally, because the minutes of social media use were positively skewed, we created a low to high quartiles of social media use variable to include in regression analyses.

### Alcohol use

collected at wave 1 and wave 3, measured the number of days in the past seven days that respondents consumed alcohol.

### Social distancing

Respondents were also asked at wave 1 and wave 3 to indicate what actions they personally took in the last seven days to keep safe from the coronavirus. We examined in the current study whether respondents **worked or studied from home** (0 = no, 1 = yes).

### Sociodemographic variables

included sex (male or female), race/ethnicity, marital status (married, separated/divorced/widowed, or never married), level of education (less than high school, high school/GED, some college, and a bachelor’s degree or higher), current employment (yes or no), and age categorized into groups (18-29, 30-44, 45-64, and 65 years or more). The race/ethnicity variable combines respondents’ answers to questions about race and about Hispanic/Latin ethnicity into five groups (non-Hispanic Asian, non-Hispanic Black, Hispanic/Latino, non-Hispanic White, and non-Hispanic individuals of other races).

### Data analysis

Analyses were conducted using Stata’s SVY command (23) to account for the UAS survey’s weight information. Descriptive statistics on all variables were generated for the study sample. Bivariate tests, *t* tests for continuous variables and chi-square for categorical variables, compared social media users on sociodemographic characteristics and on alcohol consumption and social distancing behaviors at wave 1 and wave 3. These tests looked at Twitter, Instagram, and Facebook use separately.

All multivariate model testing used linear regression with alcohol consumption at wave 3 as the outcome. Models were tested in four steps: first (model 1) to examine social media use and working/studying from home as independent variables; then (model 2) to re-examine these variables after accounting for respondents’ alcohol consumption at wave 1; and, next (model 3), after accounting for sociodemographic variables. In the model, we controlled for the selected socio-demographics after preliminary analyses (see Supplemental Table S1) showed that respondents who did and did not use Twitter, Instagram, and Facebook were different on these variables. As a final, and 4th step, we tested interactions between social media use and working/studying from home at wave 3. We added one interaction term at a time for Twitter, Instagram, and Facebook use. We plotted statistically significant social media by work/study from home interactions. A probability value of .05 was used to determine statistical significance. All reported coefficients are unstandardized.

## Results

### Social media use at wave 1

Most respondents in the sample used Facebook (67.98%), with fewer using Instagram (32.50%) or Twitter (19.61%). On average, respondents spent one hour per day on social media (*M* = 61.04, *SE* = 1.82), although this was higher for Twitter users (*M* = 100.69, *SE* = 1.58) and Instagram users (*M* = 97.799, *SE* = 4.23) than Facebook users (*M* = 80.02, *SE* = 2.43). Thirty-three percent of respondents in the current sample reported using more than one social media platform (0 platforms 26.39%; one 39.97%; two 20.85%; and three 12.80%).

### Alcohol use and social distancing behaviors at wave 1 and 3 by social media platform

Respondents reported drinking, on average, one day over the past 7 days at wave 1 and somewhat more than one day at wave 3. Alcohol consumption did not vary by social media use at wave 1, but was significantly greater at wave 3 for Twitter users and Instagram users than for non-users. There was no statistically significant difference in alcohol use at wave 3 between Facebook users and non-users. Related to social distancing, about a quarter of all respondents were either working or studying from home at wave 1, and this increased to about half of respondents at wave 3. At both wave 1 and wave 3, a larger percentage of Twitter and Instagram users compared to non-users on these platforms were working or studying from home. This was only the case for Facebook users compared to non-users at wave 3.

### Multivariate models

For model 1, we examined the relationship of respondents’ social media use at wave 1 and social distancing behavior at waves 1 and 3 to alcohol consumption at wave 3. Instagram use and time spent on social media across platforms were statistically significant in predicting consumption, but time spent had a negative correlation. Being an Instagram user was associated with greater consumption, while more time spent on social media across platforms was associated with less consumption. Working from home at wave 3, but not wave 1, was associated with greater alcohol consumption at wave 3.

For model 2, we re-evaluated these relationships after accounting for wave 1 alcohol consumption. Instagram use and time spent on social media across platforms were no longer associated, while Twitter use was now associated with greater alcohol consumption. The variance explained by working from home was reduced, but remained statistically significant and associated with greater alcohol consumption.

These relationships again changed after including sociodemographic variables in the model. Instagram use was again positively associated with greater alcohol consumption, although the variance explained was reduced. Facebook use was now associated with less alcohol consumption, and Twitter use was no longer associated. Again, the variance explained by working from home was reduced, but it remained statistically significant and positively associated with alcohol consumption.

When testing whether working/studying from home at wave 3 moderated the relationships between social media use and alcohol consumption (models not shown), the interaction between Twitter use and working/studying from home was statistically significant (*B* = .268, *SE* = .132 and *p* = 0.043) in association with wave 3 alcohol consumption. The interactions including Instagram use (*B* = .168, *SE* = .114 and *p* = 0.146) and Facebook use (*B* = .019, *SE* = .107 and *p* = 0.875) with working/studying from home were not associated. Figure 1 provides a visual representation of the Twitter use by working/studying from home interaction.

**Table 2.**
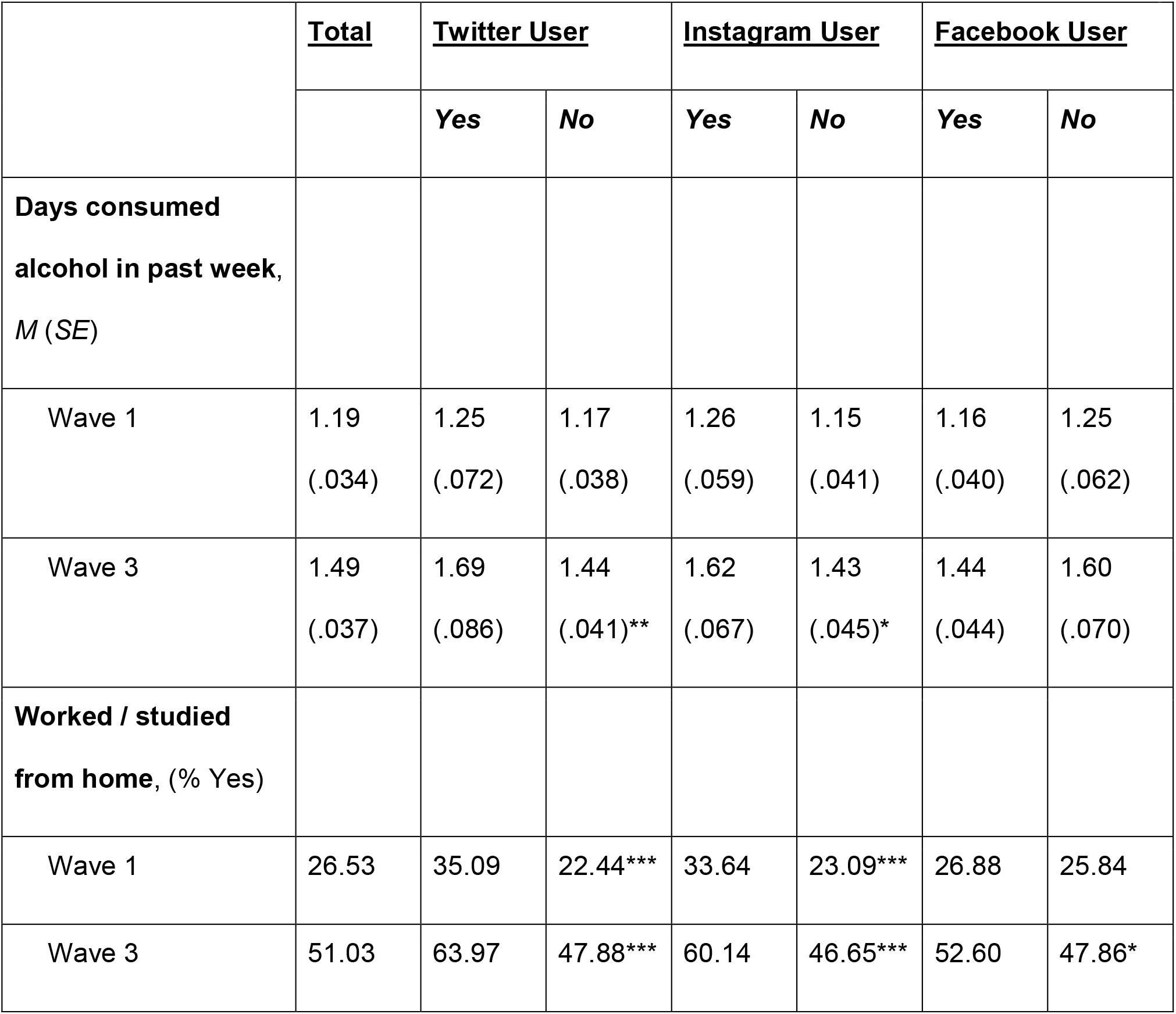
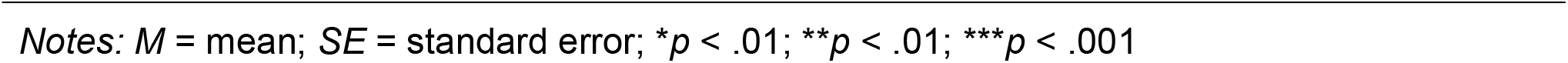
Alcohol and social distancing behaviors by social media use (*N*=5874)

|  | <u>Total</u> | <u>Twitter User</u> |  | <u>Instagram User</u> |  | <u>Facebook User</u> |  |
| --- | --- | --- | --- | --- | --- | --- | --- |
|  |  | Yes | No | Yes | No | Yes | No |
| **Days consumed alcohol in past week, *M (SE)*** |  |  |  |  |  |  |  |
| Wave 1 | 1.19<br>(.034) | 1.25<br>(.072) | 1.17<br>(.038) | 1.26<br>(.059) | 1.15<br>(.041) | 1.16<br>(.040) | 1.25<br>(.062) |
| Wave 3 | 1.49<br>(.037) | 1.69<br>(.086) | 1.44<br>(.041)** | 1.62<br>(.067) | 1.43<br>(.045)* | 1.44<br>(.044) | 1.60<br>(.070) |
| **Worked / studied from home, (% Yes)** |  |  |  |  |  |  |  |
| Wave 1 | 26.53 | 35.09 | 22.44*** | 33.64 | 23.09*** | 26.88 | 25.84 |
| Wave 3 | 51.03 | 63.97 | 47.88*** | 60.14 | 46.65*** | 52.60 | 47.86* |

Notes: $M$ = mean; $SE$ = standard error; \* $p < .01$ ; \*\* $p < .01$ ; \*\*\* $p < .001$

**Table 3.** Regression models for wave 3 alcohol consumption.

|  | **<u>Model 1</u>** | **<u>Model 2</u>** | **<u>Model 3</u>** |
| --- | --- | --- | --- |
|  | *B (SE)* | *B (SE)* | *B (SE)* |
| **Social media use** |  |  |  |
| Twitter user (%) | .198 (.104) | .153 (.072)* | .081 (.071) |
| Instagram user (%) | .256 (.091)** | .093 (.062) | .138 (.064)* |
| Facebook user (%) | -.091 (.094) | -.079 (.059) | -.130 (.058)* |
| Quartiles of minutes spent<br>on social media | -.147 (.039)*** | -.034 (.026) | -.005 (.026) |
| **Social distancing** |  |  |  |
| Worked / studied from home<br>– wave 1 | -.105 (.093) | -.103 (.065) | -.080 (.066) |
| Worked / studied from home<br>– wave 3 | .363 (.080)*** | .230 (.055)*** | .181 (.056)** |
| **Wave 1 alcohol use** |  |  |  |
| Days consumed alcohol,<br>past week | — | .838 (.017)*** | .825 (.018)*** |
| **Sociodemographic**<br>**variables** |  |  |  |
| Female (ref: Male) | — | — | -.094 (.057) |
| Age (ref: 65 +) |  |  |  |
| 18-29 | — | — | -.017 (.112) |
| 30-44 | — | — | .038 (.086) |
| 45-64 | — | — | .069 (.067) |
| Race/ethnicity (ref: White) |  |  |  |
| Asian | — | — | -.502 (.110)*** |
| Black | — | — | -.103 (.105) |
| Hispanic/ Latino | — | — | -.069 (.086) |
| Other | — | — | -.180 (.118) |
| Education (ref: Bachelor's +) |  |  |  |
| < High school | — | — | -.189 (.102) |
| High school/ GED | — | — | -.256 (.074)** |
| Some college | — | — | -.189 (.067)** |
| Employed (ref: No) | — | — | .075 (.059) |
| Marital status (ref: Never married) |  |  |  |
| Married | — | — | .161 (.076)* |
| Previously married <sup>a</sup> | — | — | .156 (.092) |
| Household members | — | — | .001 (.021) |

Notes: \* $p < .05$ ; \*\* $p < .01$ ; \*\*\* $p < .001$ ; <sup>a</sup> Separated/divorced/widowed

**Figure 1.**
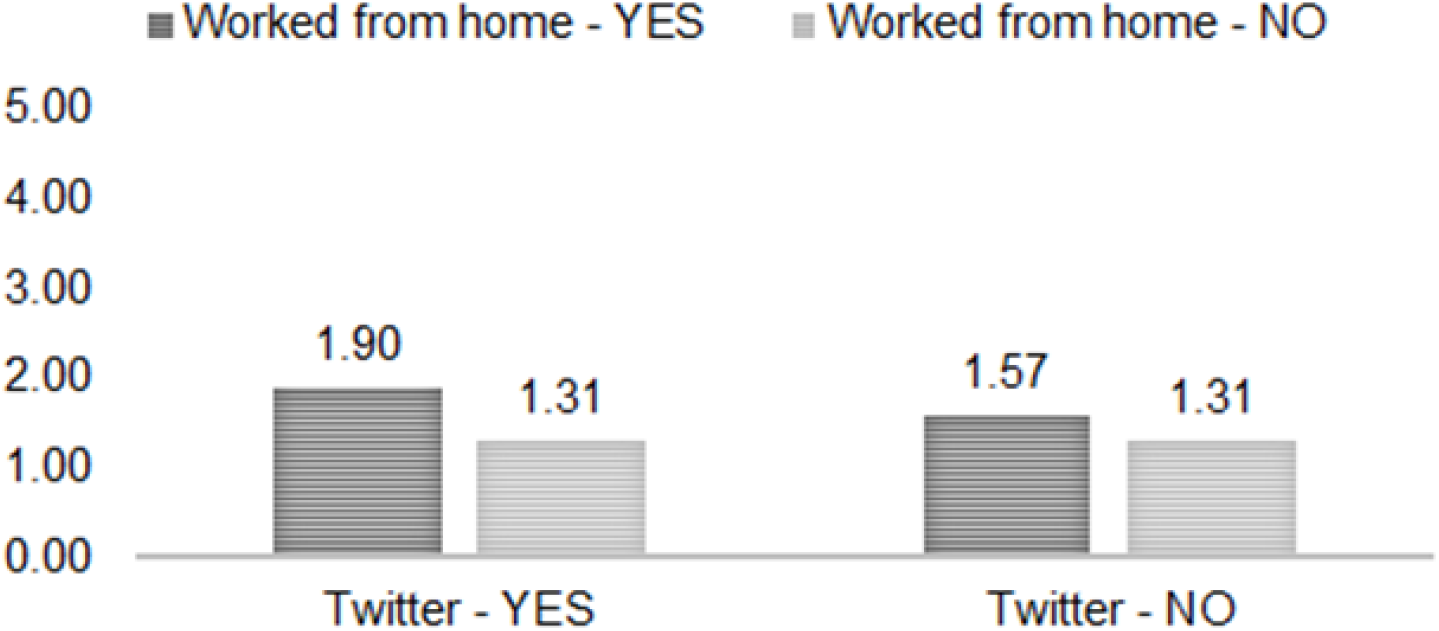
Twitter use by working from home: Mean days drinking in past week.

Respondents who worked/studied from home at wave 3 consumed more alcohol than those who did not, and this relationship was strongest for those who used Twitter.

## Discussion

We assessed in a U.S. general population sample of adults how social media use was associated with alcohol consumption during the months of March and April 2020 (UAS survey waves 1 and 3), when many individuals were sheltering at home because of the COVID-19 pandemic. With earlier studies showing increased drinking in association with traumatic events (18) and in association with greater social media exposure (14,16), we expected that social media use would be associated with greater alcohol use at wave 3. This hypothesis was supported, but not for all social media platforms. At wave 3, but not at wave 1, Twitter and Instagram users drank more alcohol than non-users. There was no difference in how often Facebook users and non-users drank alcohol in bivariate tests, and in multivariate models Facebook users drank less. As expected by Rehm et al. (24) in a commentary on anticipated alcohol consumption during the pandemic, we observed both increased and decreased alcohol consumption in the current study.

Two potential mechanisms were introduced by Rehm et al. (24) for these diverging effects, related to increased distress and decreased availability of alcohol. It could be that Twitter and Instagram users were exposed to more social media content about the COVID-19 pandemic, which Gao et al. (13) found in China was associated with a high prevalence of mental health problems. A 2017 meta-analysis (25) of experimental studies provides support for this mechanism, similarly showing a link between media exposure on disasters and other large-scale traumatic events, including via both mainstream sources and the internet, and negative psychological outcomes.

Additionally, in the media, the reports that alcohol consumption greatly increased in conjunction with stay at home orders was widely disseminated (e.g., Fulmer (15)), and, thus, individuals using Twitter and Instagram may have had greater exposure to these stories and to alcohol-related content on social media. There is an extensive body of research showing that drinking behaviors are positively influenced by descriptive norms (26), which are individuals’ perception of how much other people are drinking, and which are often overestimated (27). At least in adolescents, higher descriptive alcohol use norms mediate the positive relationship between exposure to substance-related social media content and alcohol use (28). With few exceptions (e.g., Lee et al. (29)), however, these relationships have not been widely examined among individuals at developmental stages beyond adolescence and young adulthood.

In multivariate models, the relationship between alcohol use at wave 3 and social media use changed depending on other variables included in the model. The increased frequency of alcohol consumption for Instagram users at wave 3 was at least partially attributed to differences in wave 1 drinking and, for Twitter users, partially attributed to their sociodemographic characteristics, as the effects of Instagram use and Twitter use were no longer statistically significant when these variables were added to the model. In particular, Instagram users are on average among the youngest users on social media platforms, while Facebook use is common across a range of ages and Twitter use is more common in middle-aged adults (30,31). The prevalence of risky alcohol use also trends younger, with rates of binge drinking peaking in young adulthood and decreasing thereafter (32). For demographic effects, those respondents with a bachelor’s degree or higher drank more frequently at wave 3 than those with lower levels of education. Twitter users in supplemental analyses showed the highest percentage of individuals with a bachelor’s degree or higher, which may contribute to their increased drinking at wave 3.

It was unexpected that respondents who reported more time on social media at wave 1 drank less alcohol at wave 3, although this relationship appears to also be partially attributed to how often they were drinking at wave 1. Given the lower percentage of respondents working or studying from home at wave 1 (27%-35%) compared to wave 3 (53%-64%), it could be that those individuals who were drinking more frequently at wave 1 were less likely to be at home and on social media. Working or studying from home at wave 3, but not at wave 1, was associated with greater alcohol use.

For the current study, we further expected that the positive relationship between social media use and alcohol consumption would be stronger for individuals who were working or studying from home. This hypothesis was supported for Twitter users, but not for Instagram or Facebook users. In multivariate models, the combination of working/studying from home and being on Twitter was associated with drinking more days a week. In our sample, a larger percentage of Twitter users and Instagram users than non-users were working or studying from home at wave 1 and at wave 3, and a somewhat larger percentage of Facebook users compared to non-users were working or studying from home at wave 3. It could be that this relationship was not shown for Instagram users because most of them skew younger, and young millennials and older members of Gen Z that tend to use Instagram more have increasingly been moving home with their parents because of the COVID-19 related economic downturn, where they may have less access to alcohol (33–35). Rehm et al. (24) pointed to this reduced access as being one potential mechanism influencing alcohol use during the pandemic, which, suggested by this study, may be more likely to affect younger adults than middle-aged adults.

### Strengths and limitations

This study has some unique strengths. Because of the timeliness of the UAS data collections, we were able to use current data on social media use and alcohol consumption during the early months of the COVID-19 pandemic, the depth and breadth of which was not readily available via other sources. Self-reported alcohol use and social distancing behaviors were available across two time points, which allowed for an analysis of patterns of and changes in behavior. The analysis was also completed in a weighted sample, making the results generalizable to the U.S. population; however, the focus on older groups and individuals more likely to be engaged in post-secondary education may make the results less generalizable to more vulnerable groups and those under 18 years of age, for whom social media use and alcohol consumption habits and impacts may differ.

This study is not without limitations. The UAS gathered information on the frequency of alcohol use, but not the amount consumed; as a result, it is clear how many days per week individuals self-reported consumption of alcohol, but not the overall quantity of consumption. Social media use was also not categorized by function, so different populations may have used social media for different purposes, the content of which (e.g., whether alcohol-related) was not included in the study.

### Implications for prevention

Regardless of the cause of increased alcohol use, even a moderate increase can have severe economic, health, and social implications. In the U.S., alcohol use and abuse have been attributed to significant economic losses. Sacks et al. (36) found that in 2010, alcohol use cost the U.S. approximately $249.0 billion; 72% of the costs were associated with reduced workplace productivity and 11% to healthcare expenses. Alcohol consumption has also been linked to negative short- and long-term health effects and effects on mortality measures, ranging from increases in alcohol-related cancers to accidental alcohol-related accidents (37).

Most relevant to pandemic and COVID-19 is scientific evidence that alcohol use reduces immune functioning (38), making individuals who drink more heavily more susceptible to infection. As such, the current study has implications for prevention strategies regarding COVID-19 and alcohol consumption. Social media platforms are increasingly being used for disease detection and outbreak tracking allowing epidemiologists and public health researchers rapid detection of infectious diseases and their spread (39–41), as well as for identifying at-risk drinkers and for the delivery by web-based interventions (42). For these reasons, measurements of alcohol use and social media use, as well as their correlation, may be relevant in the prevention, tracking, and treatment.

During the pandemic, and for some a concurrent “infodemic,” social media could be used as a resource to provide access to otherwise unavailable public health information, as well as peer and emotional support around health and safety. In studying social media’s use in health communication, Moorhead et al. (11) found that social media improved the tailoring, sharing, and availability of health communication. However, it was also found that much health information shared via social media lacked reliability and quality that may deter vulnerable populations from seeking care (11). Given this information, the potential reach and impact of public health messaging via social media regarding alcohol consumption should not be understated, and the application of social media campaigns (e.g., Perkins et al. (43)) that seek to influence alcohol use behaviors warrants future research under traumatic conditions.

## Data Availability

The data that support the findings of this study (identifier: UAS230 and UAS240) cannot be shared publicly without restriction. These data are accessible from the Center for Economic and Social Research, Understanding America Study (UAS), data archives at the University of Southern California to researchers who create a UAS data account and provide the appropriate signed data use agreement.

https://uasdata.usc.edu

## Acknowledgements

We would like to thank Tommy Buckley for his work in preparing this paper for submission. The project described in this paper relies on data from surveys administered by the Understanding America Study, which is maintained by the Center for Economic and Social Research (CESR) at the University of Southern California. The content of this paper is solely the responsibility of the authors and does not necessarily represent the official views of USC or UAS. The collection of the UAS COVID-19 tracking data is supported in part by the Bill & Melinda Gates Foundation and by grant U01AG054580 from the National Institute on Aging. The funders played no role in this study’s design, data analysis, the decision to publish, or the preparation of the manuscript.

## Supporting information

**S1 Table.** Sample socio-demographics by Twitter, Instagram, and Facebook use (*N*=5874)

|  | <u>Did not use</u><br><u>Twitter</u> | <u>Used</u><br><u>Twitter</u> | *p* | <u>Did not use</u><br><u>Instagram</u> | <u>Used</u><br><u>Instagram</u> | *p* | <u>Did not use</u><br><u>Facebook</u> | <u>Used</u><br><u>Facebook</u> | *p* |
| --- | --- | --- | --- | --- | --- | --- | --- | --- | --- |
| **Sex (% Female)** | 51.27 | 49.52 |  | 44.12 | 65.11 | *** | 40.08 | 56.04 | *** |
| **Age (%)** |  |  | *** |  |  | *** |  |  | *** |
| 18-29 years | 9.77 | 22.23 |  | 6.38 | 24.34 |  | 8.48 | 13.99 |  |
| 30-44 years | 22.37 | 29.58 |  | 19.57 | 32.53 |  | 24.73 | 35.61 |  |
| 45-64 years | 15.07 | 18.18 |  | 14.80 | 17.53 |  | 37.62 | 33.49 |  |
| 65 years or older | 17.81 | 14.92 |  | 19.28 | 13.02 |  | 29.18 | 16.90 |  |
| **Race/ethnicity (%)** |  |  | *** |  |  | *** |  |  | *** |
| Asian<sup>a</sup> | 5.08 | 5.97 |  | 4.47 | 6.89 |  | 6.81 | 4.53 |  |
| Black<sup>a</sup> | 12.16 | 8.73 |  | 12.43 | 9.53 |  | 14.34 | 10.16 |  |
| Hispanic / Latino | 14.2 | 20.14 |  | 12.47 | 21.37 |  | 16.44 | 14.87 |  |
| White<sup>a</sup> | 65.61 | 59.8 |  | 67.78 | 57.59 |  | 59.61 | 66.72 |  |
| Other groups<sup>a</sup> | 2.95 | 5.36 |  | 2.85 | 4.62 |  | 2.80 | 3.73 |  |
| **Education (%)** |  |  | *** |  |  | *** |  |  | ** |
| Less than high school | 9.34 | 4.72 |  | 9.87 | 5.43 |  | 10.19 | 7.54 |  |
| High school/ GED | 30.95 | 20.65 |  | 31.65 | 23.30 |  | 31.27 | 27.82 |  |
| Some college | 28.20 | 26.96 |  | 27.11 | 29.74 |  | 25.24 | 29.27 |  |
| Bachelor's degree + | 31.51 | 47.67 |  | 31.37 | 41.53 |  | 33.30 | 35.37 |  |
| **Currently employed (% Yes)** | 58.31 | 70.89 | *** | 55.61 | 71.51 | *** | 53.35 | 64.30 | *** |
| **Marital status (%)** |  |  | *** |  |  | *** |  |  | *** |
| Married | 57.11 | 55.25 |  | 58.66 | 52.77 |  | 57.60 | 56.42 |  |
| Previously married<sup>b</sup> | 20.78 | 13.14 |  | 21.73 | 14.22 |  | 20.15 | 18.78 |  |
| Never married | 22.11 | 31.61 |  | 19.61 | 33.01 |  | 22.25 | 24.81 |  |
| **Household members, *M* (*SE*)** | 1.73 (.031) | 2.03 (.068) | *** | 1.66 (.033) | 2.05 (.052) | *** | 1.57 (.048) | 1.89 (.034) | *** |

Notes: *M* = mean; *SE* = standard error; \* $p < .01$ ; \*\* $p < .01$ ; \*\*\* $p < .001$ ; GED = general educational diploma; <sup>a</sup>non-Hispanic; <sup>b</sup>separated/ divorced/ widowed

